# High frequency of bedaquiline resistance in programmatically treated drug-resistant TB patients with sustained culture-positivity in Cape Town, South Africa

**DOI:** 10.1101/2022.11.14.22282167

**Authors:** B. Derendinger, A. Dippenaar, M. de Vos, S. Huo, R. Alberts, R. Tadokera, J. Limberis, F. Sirgel, T. Dolby, C. Spies, A. Reuter, M. Folkerts, C. Allender, A. Van Rie, S. Gagneux, L. Rigouts, J. te Riele, K. Dheda, D. Engelthaler, R. Warren, J. Metcalfe, H. Cox, G. Theron

**Affiliations:** DSI-NRF Centre of Excellence for Biomedical Tuberculosis Research, SAMRC Centre for Tuberculosis Research, Division of Molecular Biology and Human Genetics, Faculty of Medicine and Health Sciences, Stellenbosch University, Cape Town, South Africa; Family Medicine and Population Health, Faculty of Medicine and Health Sciences, University of Antwerp, Antwerp, Belgium; Department of Mycobacteriology, Institute of Tropical Medicine, Antwerp, Belgium; Natera, Inc, San Carlos, USA; Division of Pulmonary and Critical Care Medicine, Zuckerberg San Francisco General Hospital and Trauma Center, University of California San Francisco, USA; National Health Laboratory Services Green Point, Cape Town, South Africa; Médecins Sans Frontières, Khayelitsha, South Africa; Translational Genomics Research Institute, Flagstaff, USA; Swiss Tropical and Public Health Institute, Allschwil, Switzerland; University of Basel, Basel, Switzerland; Brooklyn Chest Hospital, Cape Town, South Africa; Division of Pulmonology, Department of Medicine, Centre for Lung Infection and Immunity, University of Cape Town Lung Institute, Cape Town, South Africa; Centre for the Study of Antimicrobial Resistance, South African Medical Research Council; Department of Infection Biology, Faculty of Infectious and Tropical Diseases, London School of Hygiene and Tropical Medicine, London, United Kingdom; Division of Medical Microbiology, Department of Pathology, University of Cape Town, South Africa; Institute of Infectious Disease and Molecular Medicine and Wellcome Centre for Infectious Disease Research, University of Cape Town, South Africa

## Abstract

**Introduction:** Bedaquiline (BDQ) is a lifesaving new tuberculosis (TB) drug undergoing global scale-up. Data on resistance emergence in programmatic settings, especially in patients resistant to other drugs with potentially weak background regimens, is scarce. Such individuals are a priority for novel drug access yet a potential source of population-level resistance.

**Methods:** We collected culture isolates from 40 drug resistant (DR)-TB patients, culture-positive after ≥4 months of BDQ-based treatment at baseline (pre-BDQ treatment initiation) and follow-up (closest post-four-month isolate). We did MGIT960 (1μg/ml) BDQ drug susceptibility testing (DST), targeted deep sequencing (TDS; *Rv0678, atpE, pepQ*), and whole genome sequencing (WGS). Contemporaneous programmatic BDQ DST was unavailable.

**Results:** Eight percent (3/40) of patients’ strains were BDQ resistant at baseline, and 47% (19/40) gained BDQ phenotypic resistance [88% (15/17) due to acquisition, 12% (2/17) reinfection]. Several single nucleotide polymorphisms and indels in *rv0678* and *pepQ* were associated with phenotypic resistance but none in *rv0676c* and *rv1979c* (potential lineage markers). TDS detected low-level variants undetected by WGS, however, none were in genes without WGS-detected variants. Patients with baseline fluoroquinolone-resistance, clofazimine exposure, and ≤4 effective drugs were more likely to be BDQ-resistant at follow-up.

**Conclusion:** BDQ resistance acquisition, for which we identified risk factors, was common in these programmatically treated patients. Our study highlights risks associated with implementing new drugs in such populations. Likely BDQ resistance transmission occurred. Routine BDQ DST should urgently accompany scale-up of new all oral regimens, however, rapid BDQ genotypic DST remains challenging given the diversity of variants observed.

## Introduction

Tuberculosis (TB) is an ongoing health crisis. Drug-resistant (DR)-TB is difficult to diagnose and treat (1). Bedaquiline (BDQ), a novel World Health Organization (WHO)-endorsed TB drug introduced for limited compassionate use in South Africa in 2012 (2), is now a part of all standard treatments in DR-TB in South Africa (key component of injectable-free regimens) (3).

BDQ phenotypic DST (pDST) should be done before patients are started on a BDQ-containing regimen and to monitor resistance emergence (4). However, programmatic BDQ DST implementation has lagged. This is alarming given the occurrence of variants in BDQ candidate resistance genes existing before BDQ’s availability (5, 6), BDQ’s initial prioritisation for patients with high levels of resistance to other drugs (7), and BDQ-resistance emergence post-treatment cessation (8).

Unsurprisingly, BDQ resistance has been documented in Germany, China, and Pakistan, with resistance acquisition under programmatic settings at rates ranging between 3-17% (9-11). Furthermore, in South Africa, a study of a nationally representative sample of three thousand isolates from patients receiving BDQ found 7% to have phenotypic resistance; far exceeding rates in clinical trials (12-15).

Reduced BDQ susceptibility may be due to variants in the candidate resistance genes *Rv0678, atpE*, and *pepQ*. Further, mutations in *Rv0678* are associated with clofazimine (CFZ) resistance (16), and some isolates with elevated minimal inhibitory concentrations to BDQ have not demonstrated resistance-associated variants in any of these genes. The significance of other bacterial genes such as *Rv0676c* (*MmpL5*), *Rv0677c* (*MmpS5*), and *Rv1979c* is unknown (17-19). Our understanding of genotypic mechanisms of resistance is, however, far from complete.

Not only are more data on variants associated with phenotypic resistance needed, but there are, in general, limited data on BDQ-resistance emergence during treatment where multiple isolates are analysed from the same individuals. This is especially important in patients in whom resistance is more likely to emerge (e.g., in a programmatic rather than a clinical trial context). Such individuals could include, for example, those with high levels of resistance to second-line drugs who, despite being, on a population-level, a minority receiving BDQ (most patients do not receive BDQ), could inadvertently serve as a source of population-level BDQ resistance transmission (16). Such patients do not always rapidly clinically or bacteriologically respond positively: programmatic data from inpatients with second-line resistance in Cape Town, South Africa, show that 30% receiving BDQ as part of their regimens remain culture-positive after four months of treatment (20).

To address these gaps, we studied patients who received a BDQ-containing regimen under programmatic conditions and who remained culture-positive after receiving BDQ for at least four months; this patient group represents those for whom treatment is likely to be ineffective. We compared the baseline isolate preceding BDQ treatment initiation and next-available isolate after a four-month period of treatment, using whole genome sequencing (WGS), targeted deep sequencing (TDS), and phenotypic DST.

## Methods

### Study population

Adults with culture-positive pulmonary TB who received at least four months of a BDQ-containing regimen from DR-TB treatment facilities (as either in- or out-patients) in Cape Town, South Africa between 20 January 2016 and 20 November 2017 were identified (**Figure 1A**). Sputum was programmatically collected at baseline and each month to monitor treatment response per the national algorithm (21). The first available isolate from the sputum collected at or after four months of BDQ was designated the “follow-up isolate”.

**Figure 1.**
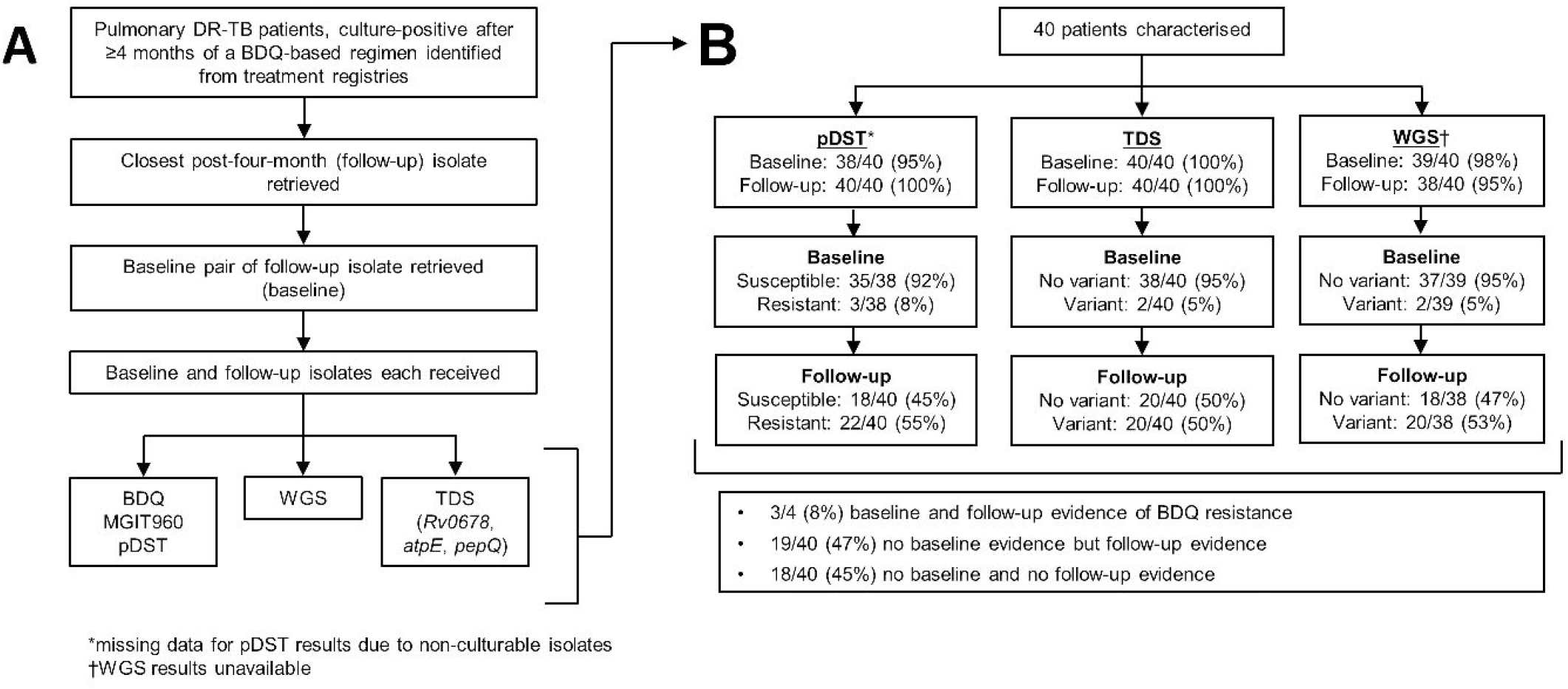
Study profile and BDQ DST results at baseline and follow-up. **(A)** Pulmonary DR-TB patients, culture-positive after _≥_4 months of a BDQ-based regimen had their baseline (close to BDQ treatment initiation) and follow-up isolate (_≥_4 months) retrieved and used for phenotypic and genotypic (TDS, WGS) drug susceptibility testing. **(B)** This approach identified a large proportion of patients whose TB isolates gained resistance: at baseline, three patients were phenotypically resistant, increasing to 22 patients at follow-up (using putative genotypic resistance markers, these numbers were 20). Only one phenotypically resistant isolate lacked genotypic evidence of resistance (follow-up, 09-A09), and all phenotypically susceptible isolates lacked variants in *Rv0678, atpE, Rv0677c*, except 1 had *pepQ* and all had *Rv0676*. variants. Abbreviations: BDQ □ bedaquiline, DR-TB □ drug resistant tuberculosis, MGIT □ mycobacterial growth indicator tube, pDST □ phenotypic drug susceptibility testing, TDS □ targeted deep sequencing, WGS □ whole genome sequencing.

Patients who had both a post-four-month isolate and a pre-BDQ treatment initiation (“baseline”) isolate stored in a biobank were included in the analysis. Our study took place in the era prior to all-oral regimens being instituted for DR-TB patients.

### Patient data collection

Patient demographic and clinical data, including previous and current TB treatment regimen and follow-up, were extracted from clinical records. Treatment outcome data were reported using WHO definitions for DR-TB contemporary with the study (22).

### BDQ phenotypic drug susceptibility testing

pDST was done on baseline and follow-up isolates in MGIT960 media as described (23). The WHO-recommended critical concentration (CC) of 1µg/ml (bedaquiline fumarate; Janssen Pharmaceuticals via the NIAID/NIH AIDS Reagent Program) was used. *Mycobacterium tuberculosis* H37Rv was used as a susceptible control and BCCM/ITM 121749 (Antwerpen, Belgium) as a resistant control (24).

### Next generation sequencing and analysis

Sequencing was done on crude or purified isolate DNA (25).

#### WGS

Genomic libraries were prepared using the DNA Prep kit (Illumina, USA) or NEBNext Ultra II kit (NEB, USA), and sequenced using NextSeq500 (Illumina) with V2, paired-end chemistry. Reads are deposited in the European Nucleotide Archive (PRJEB47429) and were analysed as described (26) to detect non-synonymous variants or indels in *Rv0678, atpE, pepQ, Rv0676c, Rv0677c*, and *Rv1979c* (19). TB profiler (version 3.0.4) was used for DST (allele frequency of _≥_10% for resistance calls). *De novo* assembly to detect *Rv0678* structural variants for one isolate with discrepant WGS and TDS results was done using UGAP https://github.com/jasonsahl/UGAP and svTyper https://github.com/SemiQuant/svTyper.git.

#### TDS

*Rv0678, atpE*, and *pepQ* were each analysed using multiple tiled-amplicons as described (8, 27) (**Supplementary Table 1**; TDS not done for other loci analysed by WGS as primers were unavailable). Samples were pooled and sequenced using an Illumina MiSeq with V3 paired-end chemistry with a targeted coverage of 30,000 reads per amplicon. Reads are in NCBI Bioproject (PRJNA767896) and were analysed by ASAP (8). Minority variants were counted if variants occurred at _≥_1% with _≥_5 paired reads.

**Table 1.**
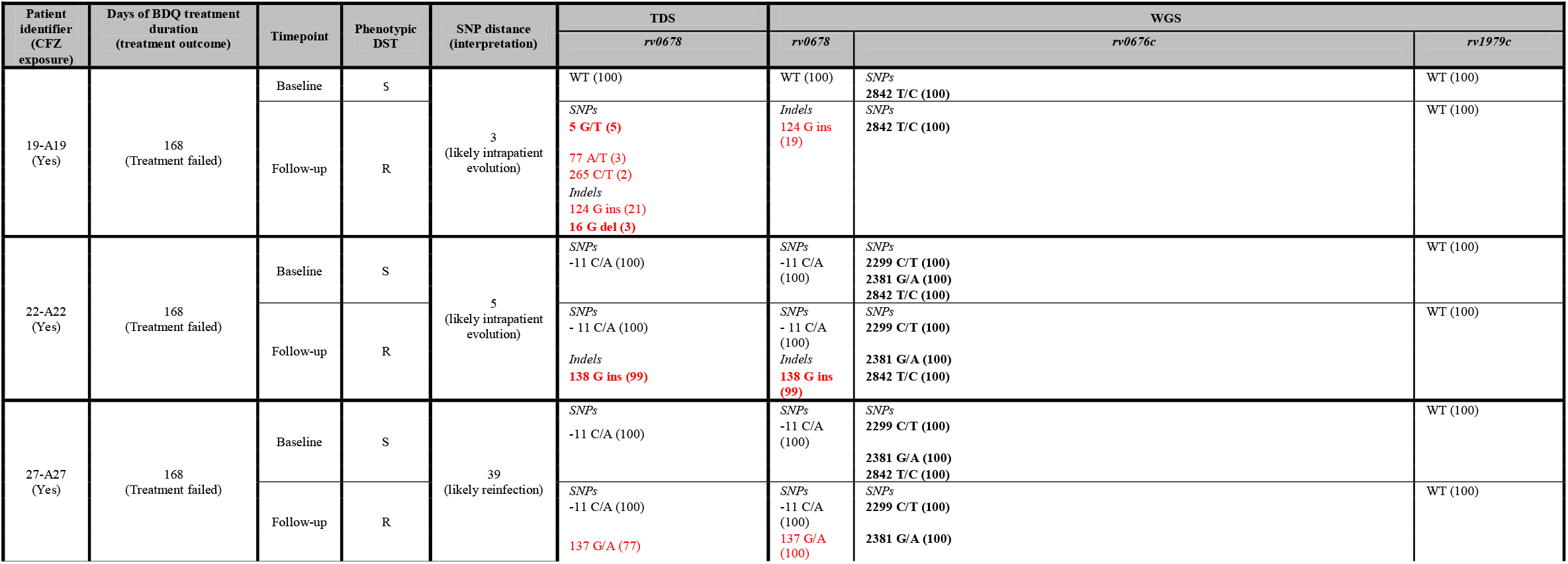
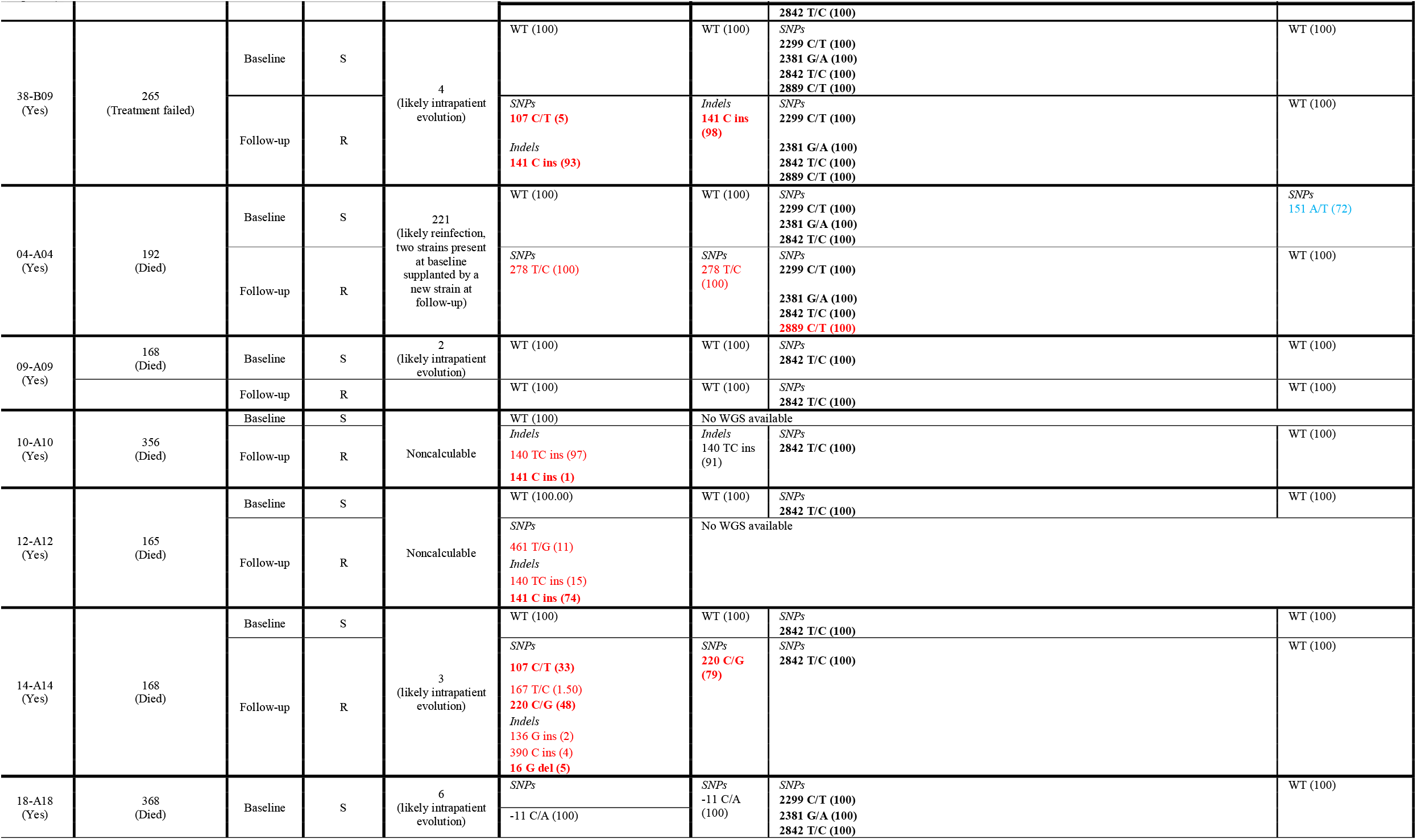
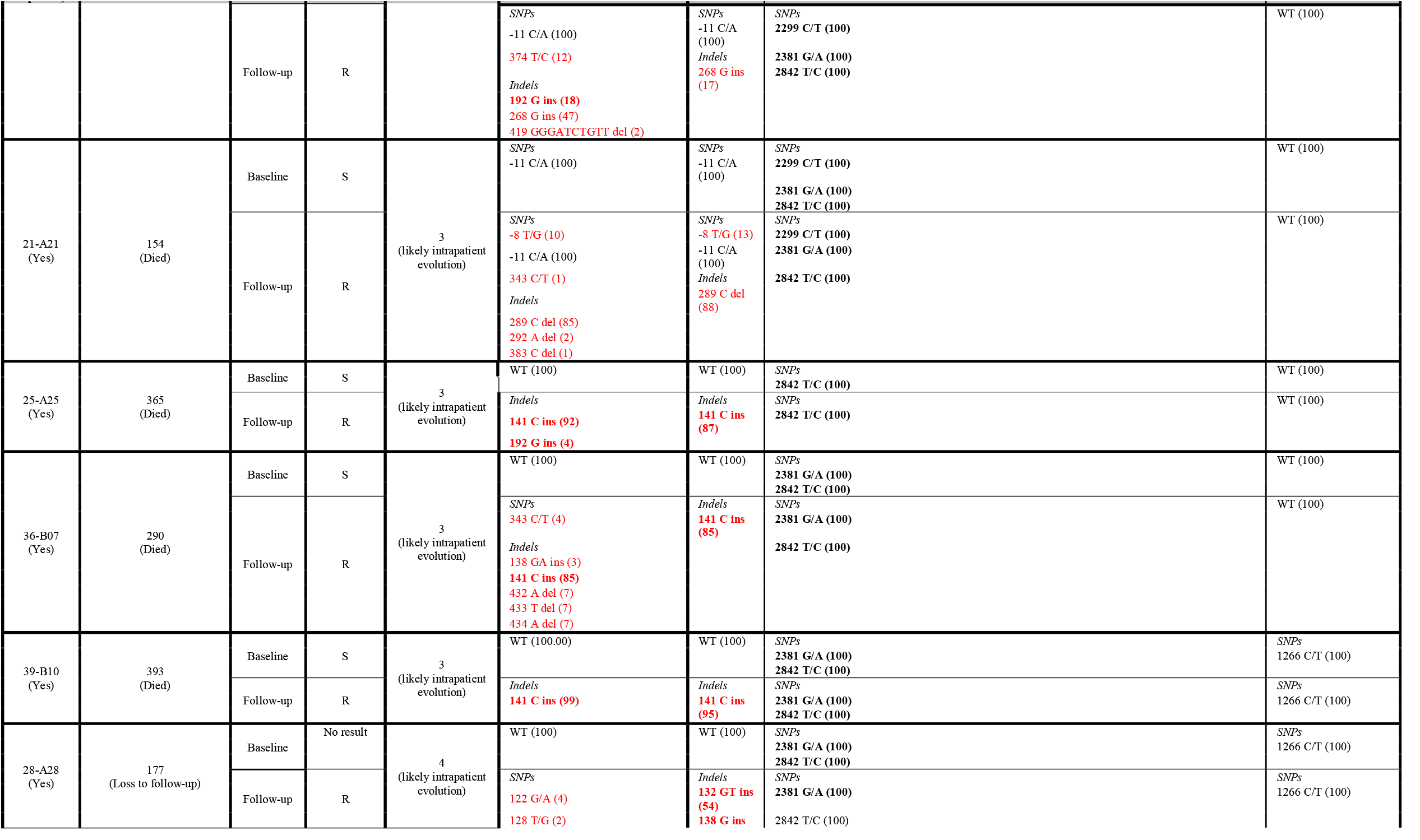
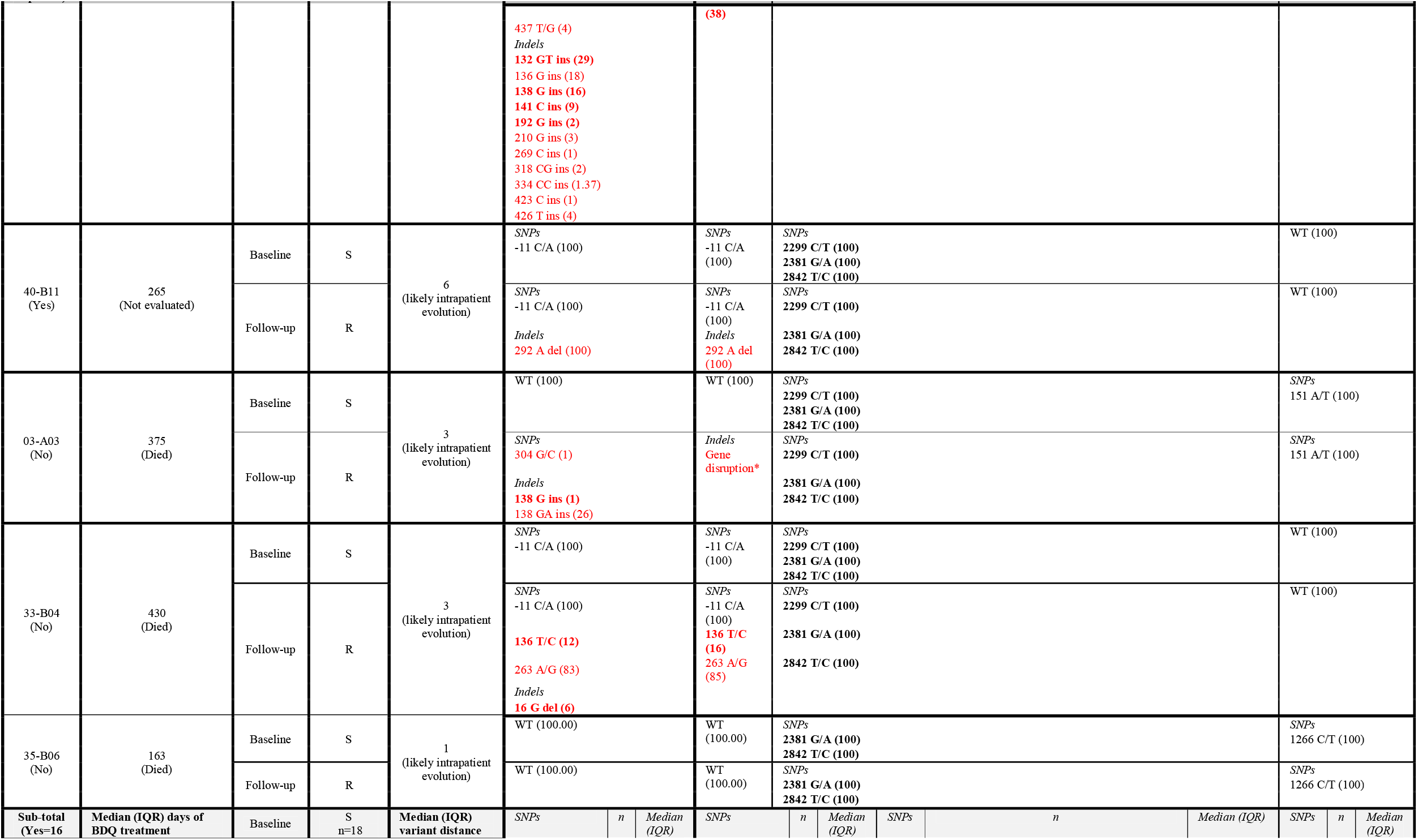
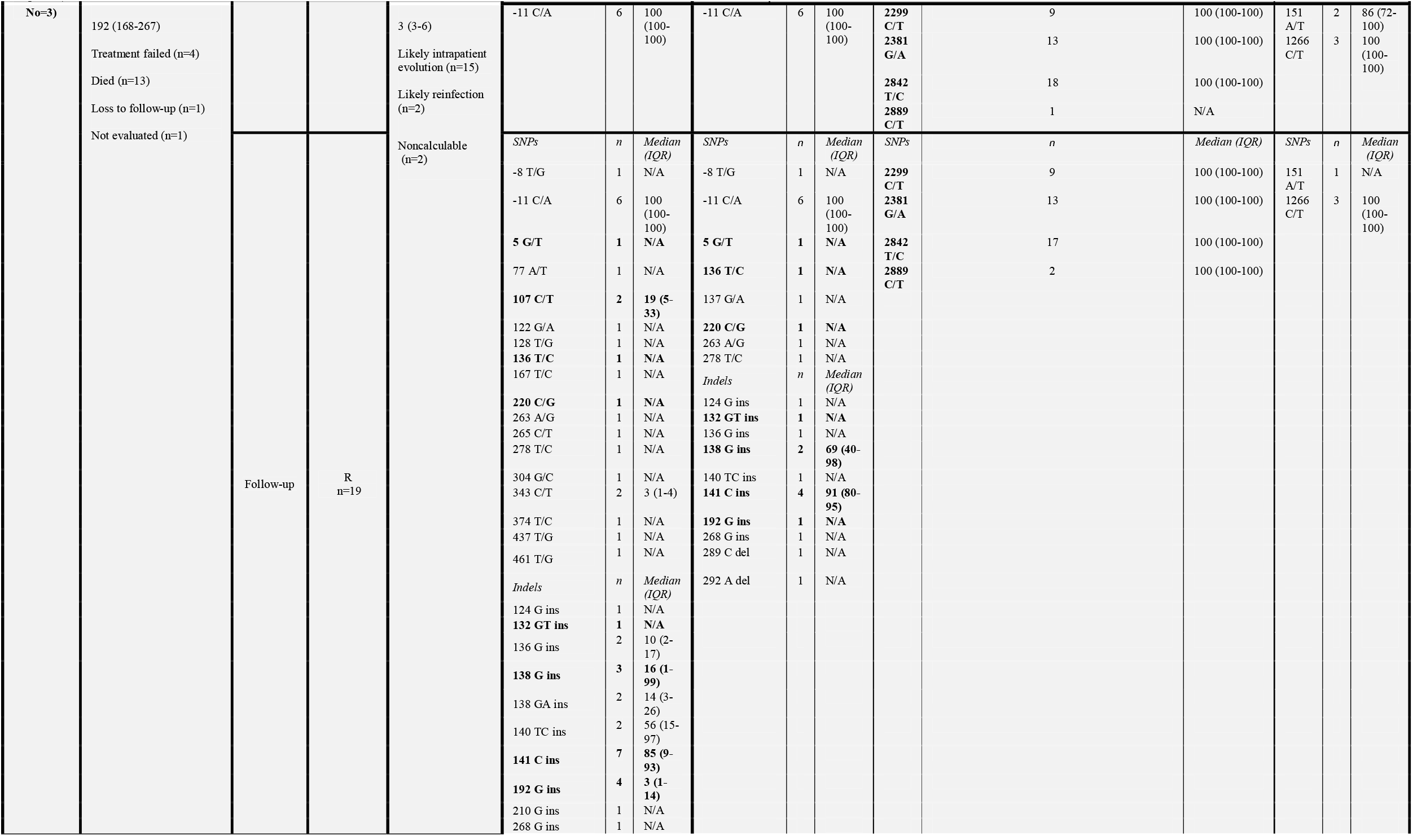
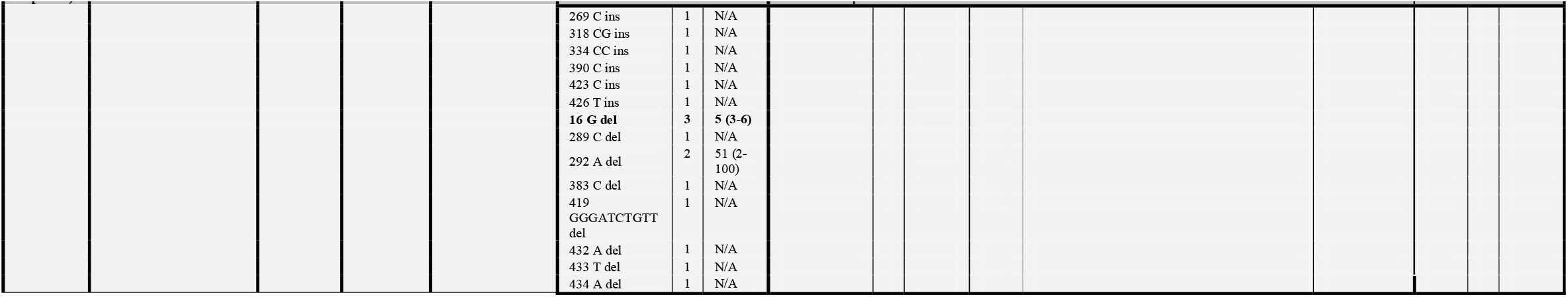
Individual patient information (phenotypic DST, TDS, WGS) in those with phenotypic BDQ resistance at follow-up but not baseline. CFZ exposure, BDQ treatment duration and outcome, whether intrapatient evolution or reinfection were likely, together with the specific variants and the percentage of reads are shown. The last row shows summary data. TDS frequently detected additional variants that WGS did not but all genes with TDS-detected variants had WGS-detected variants. *rv0678, rv0676c* and *rv1979c* variants were detected and no *atpE, pepQ* or *Rv0677c* variants were detected. Prior CFZ exposure, days on BDQ (with programmatic treatment outcome), SNP distances, and the specific variant (proportion of reads indicated) are shown (blue indicates variant loss, red indicates gain). Variants previously described(18) are bolded. In baseline susceptible isolates, most had no *rv0678* variants [33% (6/18) had -11C/A variants]. When comparing baseline and follow-up isolates, 94% (16/17) with newly gained resistance appeared to be due to intrapatient evolution, however, 6% (1/17) patients had evidence of reinfection. Abbreviations: BDQ □ bedaquiline, CFZ □ clofazimine, indels □ insertions and deletions, R □ resistant, S □ susceptible, SNPs □ single nucleotide polymorphisms, TDS □ targeted deep sequencing, WGS □ whole genome sequencing, WT □ wildtype. Data are % unless otherwise stated.

### Analyses and definitions

Statistical analyses were done using Stata (version 15; StataCorp, USA) and GraphPad Prism (version 8.0.1; GraphPad Software, USA) using 2-sided tests (_α_=0.05). McNemar’s test was used to calculate differences in paired data. For drugs other than BDQ, WGS DST was used to classify a drug as “likely effective” (Error! Reference source not found.**1** and **Supplementary Table 2**). Methods for analysing clustering, phylogeny and reinfection are in the **Supplement**. Microbiological data are in **Supplementary data file, Sheet 1**.

**Table 2.** Patient characteristics according to phenotypic BDQ resistance profile at follow-up. Those with resistance were more likely to have baseline FQ resistance, CFZ exposure, fewer likely effective drugs, and an adverse treatment outcome versus susceptible patients at follow-up. Data are n/N (%) or median (IQR). Abbreviations: ART □ antiretroviral therapy BDQ □ bedaquiline, CFZ □ clofazimine, FQ □ fluoroquinolone, HIV □ human immunodeficiency virus, IQR □ interquartile range, SLID □ second-line injectable, TB □ tuberculosis, WGS □ whole genome sequencing.

|  | Overall<br>(n=40) | BDQ phenotype at follow-up |  |
| --- | --- | --- | --- |
|  |  | Susceptible<br>(n=18) | Resistant<br>(n=22) |
| **Demographics** |  |  |  |
| Age at diagnosis of current episode (years) | 36<br>(27-45) | 34<br>(27-43) | 36<br>(27-45)<br>p=0.75 |
| Female | 11/40<br>(28) | 5/18<br>(28) | 6/22<br>(27)<br>p=0.97 |
| **Clinical characteristics** |  |  |  |
| HIV-positive | 21/40<br>(53) | 7/18<br>(39) | 14/22<br>(64)<br>p=0.12 |
| CD4 count * | 168<br>(61-394) | 235<br>(28-631) | 157<br>(66-314)<br>p=0.70 |
| ART | 18/21<br>(86) | 5/7<br>(71) | 13/14<br>(93)<br>p=0.19 |
| **Treatment history** |  |  |  |
| Previous TB | 32/40<br>(80) | 13/18<br>(72) | 19/22<br>(86)<br>p=0.27 |
| Previous drug-resistant TB | 20/32<br>(63) | 8/13<br>(62) | 12/19<br>(63)<br>p=0.93 |
| Days between baseline and follow-up isolate | 429<br>(291-564) | 369<br>(252-592) | 439<br>(365-578)<br>p=0.44 |
| Any CFZ exposure (prior or concurrent) | 29/40<br>(73) | 10/18<br>(56) | 19/22<br>(86)<br>**p=0.03** |
| Prior CFZ<sup>†</sup> | 7/26<br>(27) | 2/8<br>(25) | 5/18<br>(28)<br>p=0.88 |
| Concurrent with BDQ<sup>†</sup> | 20/26<br>(77) | 6/8<br>(75) | 14/18<br>(78)<br>p=0.88 |
| Treatment and drug resistance |  |  |  |
| BDQ treatment duration (days) | 185<br>(168-265) | 181<br>(168-267) | 193<br>(168-266)<br>p=0.80 |
| Baseline FQ resistance<sup>‡</sup> | 18/39<br>(46) | 4/18<br>(22) | 14/21<br>(67)<br>**p=0.01** |
| Overall drug resistance patient categorisation |  |  |  |
| Pre-MDR/RIF-mono | 2/36<br>(6) | 2/16<br>(13) | 0/20<br>(0)<br>p=0.10 |
| MDR | 8/36<br>(22) | 6/16<br>(38) | 2/20<br>(10)<br>**p=0.05** |
| MDR plus resistance to a FQ | 20/36<br>(56) | 7/16<br>(44) | 13/20<br>(65)<br>p=0.20 |
| MDR plus resistance to a FQ and SLID | 6/36<br>(17) | 1/16<br>(17) | 5/20<br>(25)<br>p=0.13 |
| Treatment regimen |  |  |  |
| Total number drugs (excluding BDQ)<sup>§</sup> | 7<br>(6-9) | 7<br>(6-12) | 8<br>(6-9)<br>p=0.85 |
| Likely effective<sup>¥</sup> | 4<br>(3-5) | 5<br>(4-6) | 3<br>(3-4)<br>**p<0.01** |
| Treatment outcomes |  |  |  |
| Favourable outcome | 8/40<br>(20) | 7/18<br>(39) | 1/22<br>(5)<br>**p<0.01** |
| Cured | 4/40<br>(10) | 3/18<br>(17) | 1/22<br>(5)<br>p=0.20 |
| Treatment completed | 4/40<br>(10) | 4/18<br>(22) | 0/22<br>(0)<br>**p=0.02** |
| Unfavourable outcome | 25/40<br>(63) | 6/18<br>(33) | 19/22<br>(86)<br>**p<0.01** |
| Treatment failed | 5/40<br>(13) | 0/18<br>(0) | 5/22<br>(22)<br>**p=0.03** |
| Died | 20/40<br>(50) | 6/18<br>(33) | 14/22<br>(64)<br>p=0.06 |
| **Loss to follow-up** | 4/40<br>(10) | 3/18<br>(17) | 461<br>1/22<br>(5)<br>p=0.20 |
| **Not evaluable** | 3/40<br>(8) | 2/18<br>(11) | 462<br>1/22<br>(5)<br>p=0.43 |

\* One unknown CD4 count
† Three patients had a record of CFZ treatment (two BDQ-susceptibles, one resistant) but no date
‡ Detected by WGS or programmatic line probe assay. One result unavailable
§ Two patients were excluded due to unknown background TB drug regimens
¥ Two WGS sequencing results unavailable

### Ethics

This study was approved by the Stellenbosch University Health Research Ethics Committee (N09/11/296; N16/04/045), Western Cape Health Research Committee (WC_2016RP18_637), University of California San Francisco Human Research Protection Program (14-15090) and the University of Cape Town Human Research Ethics Committee (416/2014). Patient identifiers were not known to anyone outside of the research group and cannot be used to identify a particular patient.

## Results

### Study participants

Of 40 total eligible patients (**Figure 1**), 26 (65%) were inpatients (Brooklyn Chest Hospital and Khayelitsha Day Hospital) and 14 (35%) outpatients at surrounding facilities. Twenty-eight patients (70%) were initiated on BDQ (21) when substituted for a second-line injectable (SLI) due to toxicity or intolerance, or within an individualised regimen where resistance to fluoroquinolones (FQs) and/or SLIs was noted. The remaining 12 (30%) patients were treated with BDQ-containing salvage regimens in the setting of extensive drug resistance (eight had resistance to both FQs and SLIDs, and four had histories of second-line treatment failure) (28).

### Patient and regimen characteristics

Demographics and clinical characteristics, including treatment histories and outcomes are in **Table 2**. Overall, 46% (18/39, one result unavailable) of patients had baseline WGS-detected FQ resistance, and this was more frequent in patients with BDQ resistance (at baseline and/ or follow-up) than in those who were BDQ-susceptible throughout [67% (14/21) vs. 22% (4/18); p=0.01]. Most (73%, 29/40) patients had a history of CFZ exposure and 50% (20/40) had concurrent CFZ and BDQ treatment. At follow-up, patients received a median (IQR) of 7 (6-9) drugs, and in patients who had BDQ-resistant TB, the number “likely effective” drugs were fewer than in patients with BDQ-susceptible TB [3 (3-4) vs. 5 (4-6); p<0.01]. The odds ratios (ORs, 95% CI) for BDQ resistance at follow-up for baseline FQ-resistance, CFZ exposure, and _≤_4 likely effective drugs were 7 (2-29; p<0.01), 5 (1-23 p=0.03), and 12 (2-61; p<0.01), respectively (when patients with baseline BDQ resistance and reinfection were omitted, _≤_4 likely effective drugs was the only variable with a significant OR, **Supplementary Table 3**). Levofloxacin (LFX), pyrazinamide (PZA), and CFZ (**Table 1**) were each more likely to be given ineffectively in BDQ-resistant than -susceptible patients (**Supplementary Table 2**). Patients who had phenotypically BDQ resistant isolates at follow-up had, at specimen collection, received BDQ for a similar period as susceptible patients. Over half [63% (25/40)] of all patients had an unfavourable outcome, and all but one (patient 37-B08) with a BDQ resistance-associated variant had an unfavourable outcome.

### BDQ resistance

#### Phenotypic resistance

Of baseline isolates assessable by pDST, 92% (35/38) were susceptible and 8% (3/38) resistant. Of the follow-up isolates, 45% (18/40) were susceptible and 55% (22/40) resistant. Overall, 8% (3/38) therefore had primary resistance (**Supplementary Table 4**), 47% (18/38) gained resistance (acquired or reinfection) and 45% (17/38) were susceptible at both baseline and follow-up (**Figure 1B, Figure 2**).

**Figure 2.**
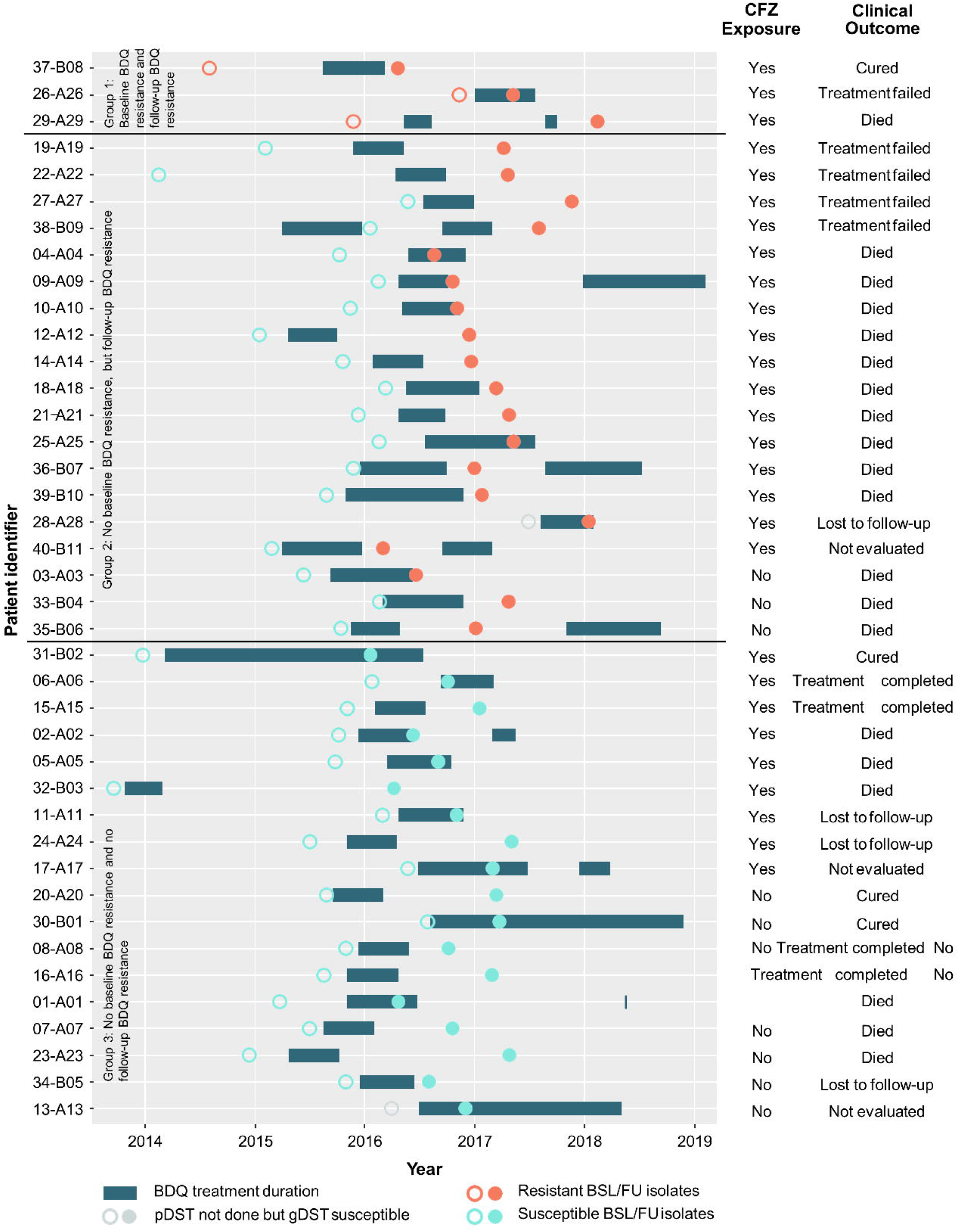
Timeline of bedaquiline treatment and follow-up isolate sampling per patient. Three patients were phenotypically resistant at baseline and follow-up (classified as Group 1), and all had CFZ exposure (prior or concurrent with BDQ). Nineteen patients acquired BDQ resistance during treatment (Group 2); all but three had CFZ exposure and all with a known treatment outcome had an unfavourable outcome. Patients susceptible at both time points were in Group 3 (n=18). Circle colouring per pDST. Abbreviations: BDQ □ bedaquiline, BSL □ baseline, CFZ □ clofazimine, FU □ follow-up, gDST □ genotypic drug susceptibility testing, pDST □ phenotypic drug susceptibility testing.

#### WGS

WGS data was available for 98% (39/40) of baseline and 95% (38/40) of follow-up isolates. In patients whose isolates gained resistance, no baseline isolates (0/17) had *Rv0678, atpE, pepQ* or *Rv0677c* variants (excluding *Rv0678* -11 C/A), though 29% (5/17) had _≥_1 *Rv1979c* variant (**Table 1**). All isolates (resistant or susceptible) had _≥_1 *Rv0676c* variant. At follow-up, the number patients whose isolates that gained variants associated with resistance increased to 88% (15/17; p<0.001 vs. baseline) for *Rv0678* and 24% (4/17; p=0.70) for *Rv1979c* (no *atpE, pepQ*, or *Rv0677c* variants were noted at follow-up). Notably, two patients’ isolates gained BDQ resistance (03-A03, 09-A09) but had no apparent variants when a genome-wide screen was done. However, after visual inspectionan a IS*6110* insertion site was identified in 03-A03. In contrast, amongst patients whose isolates remained susceptible, new variants were noted only in *pepQ* [13% (2/16), p=0.13] and *Rv1979c* [31% (5/16), p=0.69] (**Supplementary Table 5**).

#### TDS

TDS detected additional variants relative to WGS; 33% (13/40) of patient isolates with a WGS-detected variant had additional variants exclusively detected by TDS. In all patients, *Rv0678* and *pepQ* variants (none in combination) were detected in 5% (2/40) of baseline and 50% (20/40) of follow-up isolates, respectively (no *atpE* variants detected; -11C/A *Rv0678* excluded). However, no variants exclusively detected by TDS explained phenotypic BDQ resistance not otherwise explained by a WGS-detected variants. In those who gained phenotypic resistance, all but two patients [89% (16/18)] gained _≥_1 *Rv0678* variant (−11C/A *Rv0678* variants excluded) (**Table 1**). The IS*6110* insertion in patient 03-A03 follow-up isolate was undetected by TDS.

### Pairwise variant distances

Most [81% (29/36)] patients with WGS data available at baseline and follow-up had _≤_7 variants difference between isolates [median (IQR) 3 (2.5-3.7)], indicative of likely within-patient evolution (the number of variants difference was not associated with days-between-isolates, **Supplementary Figure 1**). Of these 29, 52% (n=15) transitioned phenotypically from BDQ susceptible to resistant, indicating likely resistance acquisition. The other 19% (7/36) had between 39 and 1,271 variants different, suggestive of reinfection likely from transmission. Twenty-nine percent (2/7) of these were initially BDQ susceptible and hence likely gained resistance via reinfection. Of the seven patients with likely reinfection, all but two who remained L2.2.1 (39 and 240 variants different, respectively) had a different sub-lineage at follow-up compared to baseline (**Figure 3**). Interestingly, 04-A04 had at baseline a mixed infection of two strains, both of which, based on inspection of *pncA* sequences, were not present at follow-up; rather a third new strain was present.

**Figure 3.**
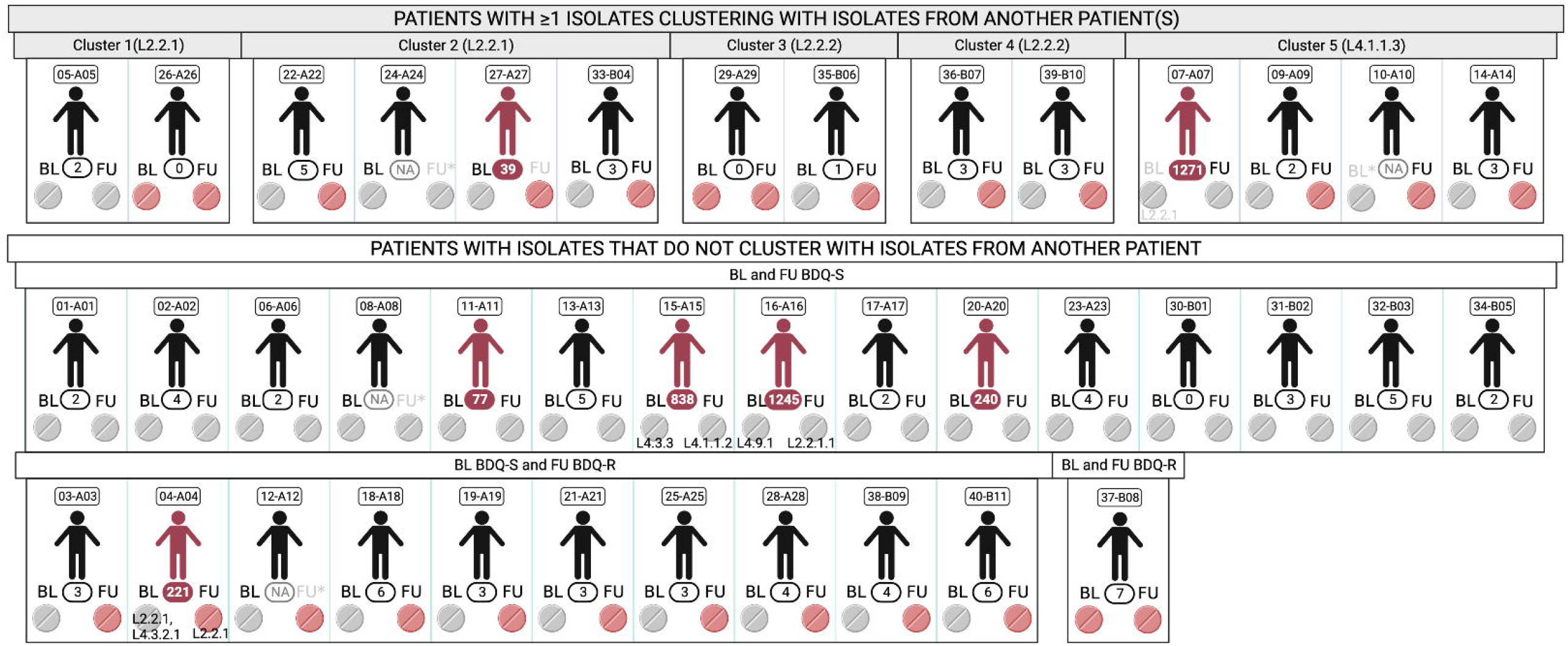
WGS of baseline and follow-up isolates identified five clusters (top row; _≤_12 SNPs) and evidence of reinfection, overlayed with phenotypic BDQ resistance statuses. Patient identifiers are above each patient with variant distances between baseline and follow-up in ovals (if distances are _≥_39, reinfection is indicated by a red patient figure and red oval background with white text). Greyed BL or FU labels indicate missing WGS data (asterisk), or the corresponding isolate does not cluster with its pair (no asterisk). Patients without isolates that cluster with other patients are grouped per baseline and follow-up phenotypic BDQ statuses (bottom two rows). Lineages are shown when pairs differed. Pill colours indicate phenotypic BDQ susceptibility (grey) or resistance (red). Abbreviations: BDQ □ bedaquiline, BDQ-R □ bedaquiline-resistant, BDQ-S □ bedaquiline-susceptible, BL □ baseline, FU □ follow-up, ID □ identification, L □ lineage, WGS □ whole genome sequencing.

### Clustering analysis

At the _≤_12 SNP cut-off, five clusters totalling 14 patients with L2 or L4 strains were identified (**Figures 3-4)**. Sixty-five percent (26/40) of patients did not cluster with one another (the _≤_5 SNP threshold identified no clusters). All clusters except Cluster 1 had at least one patient who gained phenotypic BDQ resistance and, in both Clusters 2 and 5, one patient who gained resistance had a new follow-up strain (reinfection).

**Figure 4.**
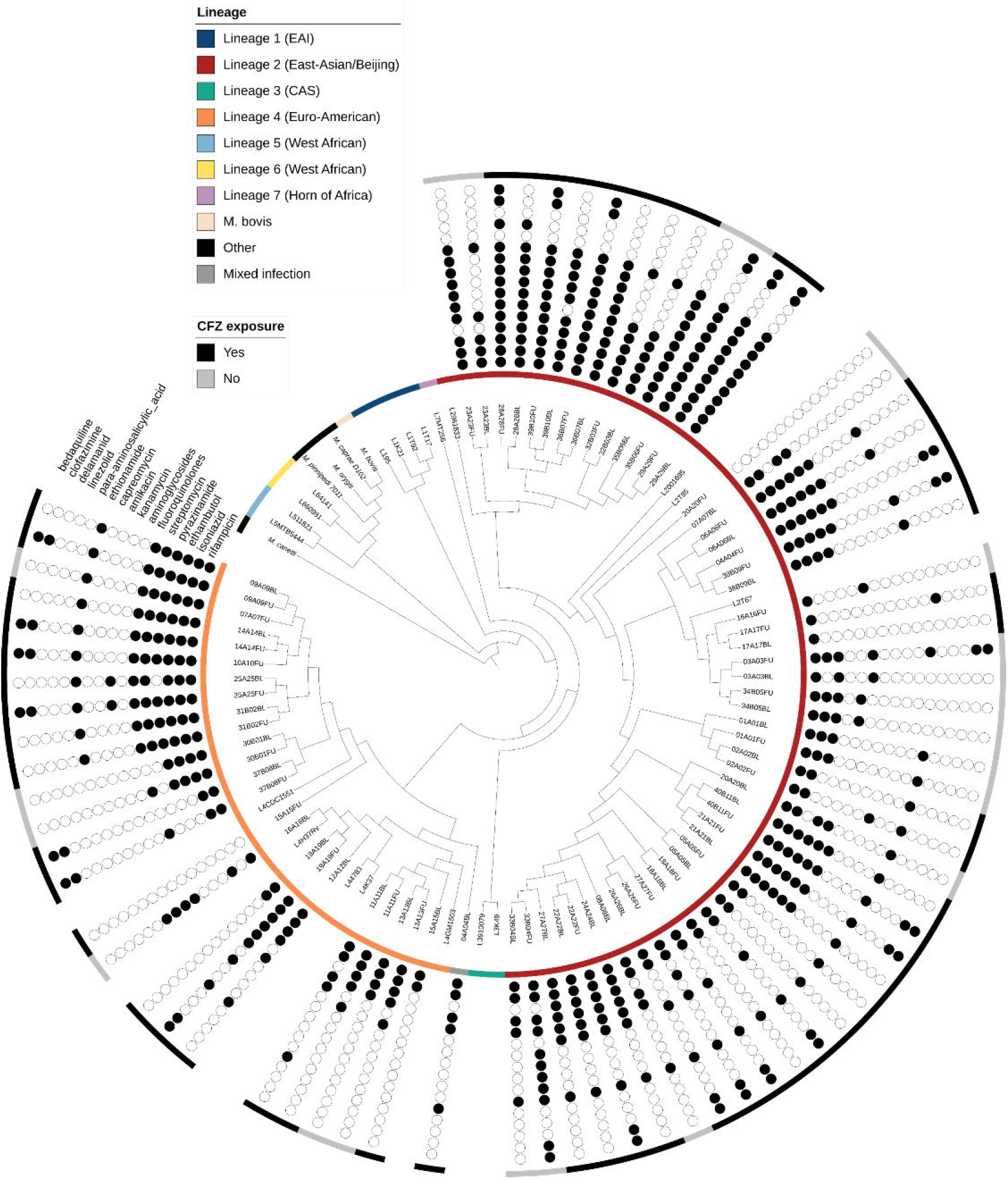
Cladogram showing lineage diversity (inner coloured ring), likely reinfection (shaded isolate identifiers), drug susceptibility (circles, based on WGS except pDST for BDQ) and CFZ exposure (outer ring). Isolates were in L2 [67% (51/76), red] or 4 [32% (24/76), orange] (one mixed with both lineages present at baseline) and isolates were resistant to many drugs. Branch lengths were ignored for visualisation and the phylogenetic analysis included 25 sequences representative of the *Mycobacterium tuberculosis* complex **(Supplementary data file, Sheet 2)**. Abbreviations: BDQ □ bedaquiline, BL□ baseline, CAS □ Central Asian, CFZ □ clofazimine, EAI □ East African Indian, FU □ follow-up, L □ lineage, pDST □ phenotypic drug susceptibility testing, WGS □ whole genome sequencing.

## Discussion

We evaluated phenotypic and genotypic BDQ resistance among programmatically treated DR-TB patients, culture-positive after four months of a BDQ-containing regimen. Key findings include: 1) more than half of the patients had isolates that gained BDQ resistance (most due to acquisition, however, reinfection may have also occurred), 2) diverse *Rv0678* and *pepQ* single nucleotide variants and indels were frequently seen with phenotypic resistance, whereas isolates with *Rv0676c, Rv0677c* and *Rv1979c* variants were seen with both susceptible and resistant isolates (suggestive of lineage markers) and no *atpE* variants were found, and 3) many minor variants were only detected by TDS, however, all isolates with exclusively TDS-detected variants already had other WGS-detected variants that were seen with phenotypic BDQ-resistance, and 4) BDQ resistance gain at follow-up was associated with fewer likely effective drugs, CFZ exposure, and baseline FQ resistance. Together, these data identify a potential programmatic source of BDQ resistance, show how reinfection can be responsible for resistance and highlight the complexity of associating specific variants with phenotypic BDQ resistance. Furthermore, these data can contribute to the WHO DR-TB mutation catalogue and provide information on which patients are most at risk of gaining resistance.

This is one of the first studies to report individual level BDQ resistance gain over time amongst patients treated in a programmatic setting with a BDQ-containing regimen. This rate was higher than described elsewhere (18, 29-31) because our patient population was deliberately pre-selected based on elevated resistance acquisition risk, and selected for likely failure of treatment. However, these individuals reflect the type of patient originally prioritised for BDQ access in our setting and, should BDQ resistance transmission become endemic, is one likely initial source (importantly, three patients were already resistant at BDQ treatment initiation). Resistance gain was primarily due to acquisition, however, in some patients it may have been due to reinfection; a finding similar to that previously described in Moldova (32).

We identified many variants in *Rv0678* and *pepQ* alongside phenotypic BDQ resistance (e.g., *Rv0678*: 343 C/T, 136 G ins, 292 A del; *pepQ:* 693 A ins) hitherto undescribed (17, 18). *Rv0676c* variants (2299 C/T, 2381 G/A, 2842 T/C) were found in both resistant and susceptible isolates at both timepoints. In the recent WHO DR-TB mutation catalogue, these variants are indeed classified as not associated with BDQ resistance (19) and are suggestive of lineage markers. Interestingly, -11C/A variants have been associated with increased BDQ susceptibility (33), however, eight of these isolates also had additional *Rv0678* variants, suggesting that any hyper-susceptibility is likely overcome.

These diverse variants represent complexity for molecular diagnostic developers that is compounded by the insertion of large elements like IS*6110* (31) in *Rv0678* that may be missed by TDS and WGS (34). This is somewhat offset by the relatively low frequency of *pepQ* variants in the absence of *Rv0678* variants (only one of these strains with only a *pepQ* variant was BDQ resistant) and a complete lack of *atpE* variants.

TDS detected many minor variants missed by WGS; however, the overall genotypic classification of resistance did not change because all variants exclusively detected by TDS were in loci that had WGS-detected variants. This may be because this patient population has a very low number of effective drugs and thus many variants can emerge – some of these to a high level (i.e., non-minority heteroresistance) at which point they are WGS-detectable. *Rv0678* may also be undergoing selection for additional variants, some of which are only detected by TDS. However, this should not rule-out the use of TDS in this population for BDQ as TDS-exclusively detected variants may affect minimum inhibitory concentrations. TDS is also useful (and often easier to do prospectively) for second-line genotypic DST (35, 36).

Several characteristics differed at baseline in patients who later gained BDQ resistance vs. those that did not. In addition to previously-described prior or concurrent CFZ exposure (37), a weaker background regimen (especially FQs being likely ineffective), puts patients at risk of resistance acquisition. This suggests that protecting the FQ (e.g., via rapid DST access or enhanced dosing) is crucial for preventing BDQ resistance acquisition, as well as ensuring that the number of likely effective drugs always exceeds four. Lastly, almost all patients with BDQ resistance had a poor clinical outcome, indicating the need for new treatment strategies in patients with BDQ-resistant TB.

Our study has strengths and limitations. First, we aimed to understand BDQ resistance emergence in a programmatic setting where we expected it to be most likely. This is intentionally not representative of the majority of patients who receive BDQ for DR-TB, for which excellent outcomes have been observed (16). Our study took place before all-oral regimens were instituted for MDR/RR-TB patients but was in a facility where novel drugs like BDQ are first used prior to population-level scale-up. On a technical level, a mix of crude or purified isolate DNA was used for sequencing, which may partly explain some discordance (e.g., TDS missing a few WGS-detected variants). Our clustering analysis was a convenient secondary analysis done to rule-in not rule-out transmission; we therefore likely underestimated transmission events.

In conclusion, this study highlights the existence of a potentially infectious pool of BDQ resistance strains, for which we show transmission occurring, created under programmatic conditions in a population of patients prioritised at the beginning of BDQ roll-out in South Africa. The study illustrates high rates of resistance in people with a delayed bacteriological response, the risks of starting patients with complex TB-treatment histories on a regimen containing a novel drug without routinely available DST (which requires balancing with ethical considerations), provides information on novel BDQ resistance- and susceptibility-associated variants, and informs upon clinical risk factors associated with resistance gain.

## Data Availability

All data produced in the present study are available upon reasonable request to the authors

## Acknowledgments

The authors thank National Health Laboratory Services, Green Point, Cape Town, South Africa. We want to thank additional members of the TGen team, including Andrew Goedderz, Jason Agundez, Meagan Papineau, Cassidy Danbury, and Darrin Lemmer for performing targeted sequencing and data analysis support. Lastly, we would like to thank the patients, their families and the healthcare staff that care for them.

## Notes

**Funding:** This work was supported by the Doris Duke Charitable Foundation (Metcalfe), the U.S. National Institute of Allergy and Infectious Diseases (NIAID) (R01AI131939, Metcalfe and Engelthaler), the South African Medical Research Council (SAMRC), National Research Foundation (NRF), the Research Foundation Flanders (FWO Odysseus, G0F8316N, Van Rie), the Stellenbosch University Faculty of Medicine Health Sciences, a Swiss-South Africa Joint Research Award (Reference: 107799, South African National Research Foundation and the Swiss National Science Foundation, Cox and Gagneux), and a Wellcome Trust Fellowship (Cox: 099818/Z/12/Z).

### Competing Interest Statement

The authors have declared no competing interest.

### Funding Statement

This work was supported by the Doris Duke Charitable Foundation (Metcalfe), the U.S. National Institute of Allergy and Infectious Diseases (NIAID) (R01AI131939, Metcalfe and Engelthaler), the South African Medical Research Council (SAMRC), National Research Foundation (NRF), the Research Foundation Flanders (FWO Odysseus, G0F8316N, Van Rie), the Stellenbosch University Faculty of Medicine Health Sciences, a Swiss-South Africa Joint Research Award (Reference: 107799, South African National Research Foundation and the Swiss National Science Foundation, Cox and Gagneux), and a Wellcome Trust Fellowship (Cox: 099818/Z/12/Z).

### Author Declarations

This study was approved by the Stellenbosch University Health Research Ethics Committee (N09/11/296; N16/04/045), Western Cape Health Research Committee (WC_2016RP18_637), University of California San Francisco Human Research Protection Program (14-15090) and the University of Cape Town Human Research Ethics Committee (416/2014).

