## Supplement for "High frequency of bedaquiline resistance in programmatically treated drug-resistant TB patients with sustained culture-positivity in Cape Town, South Africa"

### Methods

*Clustering and phylogenetic analyses*

Phylogenetic analyses included concatenated sequences of 101 *Mycobacterium tuberculosis* complex isolates (76 from the study, 25 representative of the complex (1-4) (**Supplementary data file, Sheet 2**). High confidence single nucleotide polymorphisms (SNPs) with ≥95% frequency (excluding *pe/ppe*, repeat, insertion sequences, and bacteriophages regions) (n=27,251) were used to construct a maximum likelihood phylogeny tree with IQ-TREE (version 2.0.6; 1,000 bootstrap pseudo-replicates, ultrafast automatic model selection) (5) and visualised and annotated using interactive Tree of Life (iTOL) (v6) (6). Transmission clusters were evaluated using *ape (7)* and *adegenet (8)* under SNPs thresholds of 5 and 12, with 12 (previously defined as the upper threshold of genomic relatedness noted within patient and between epidemiologically linked cases (9)) used to define a cluster. Pairwise comparisons of SNPs and indels (≤50 bp, >70% heterogeneity frequency (10)) were done between baseline and follow-up isolates of each patient to determine intrapatient variant distance.

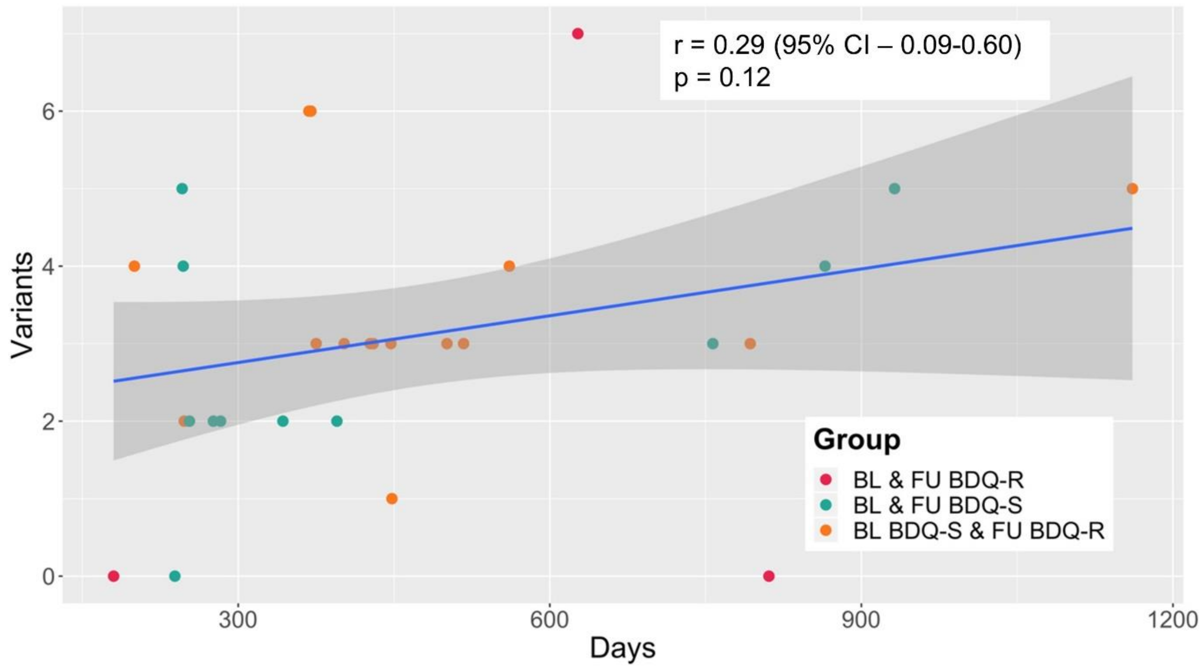

**Supplementary Figure 1.**

**Variant distance (SNPs, indels) from 29 patients showed a trend towards a positive linear correlation with days between baseline and follow-up sample collection.** Patients (n=7) with isolates with variant distances indicative of reinfection (≥39 variants apart) were omitted. Abbreviations: BDQ-S – bedaquiline susceptible, BDQ-R – bedaquiline resistant, BL – baseline, FU – follow-up, SNP – single nucleotide polymorphism.

**Supplementary Table 1.**

**BDQ resistance-associated genomic regions analysed by TDS.** Universal tail sequences (in bold) are described previously (11). Amplicons sizes and positions are given. Abbreviations: BDQ – bedaquiline, bp – base pair, F – forward, R – reverse, TDS – targeted deep sequencing

| Primer name (direction) | Sequence |
|---|---|
| ***atpE* (246 bp)** -56 bp upstream of gene to 95 bp downstream of gene** | |
| atpEf-84 (F) | **ACCCAACTGAATGGAGC**AGCCAAGCGATGGAGCTCGAAGAGGAAC |
| atpEr222 (R) | **ACGCACTTGACTTGTCTTC**GAACAGCGCCATAAAMGCCAGGTTGATG |
| atpEf130 (F) | **ACCCAACTGAATGGAGC**AGGCGCAAGGGCGGCTGTTCACA |
| atpEr+96 (R) | **ACGCACTTGACTTGTCTTC**GCTGGACTCGCCGCCTTCCTCTGC |
| ***Rv0678* (498 bp)** -33 bp upstream of gene to 495 bp in gene** | |
| Rv0678f-57 (F) | **ACCCAACTGAATGGAGC**CACGCCGGTCTGGTGACGCATACC |
| Rv0678r124 (R) | **ACGCACTTGACTTGTCTTC**AGCCCAACAATCGACCCGCCAACC |
| Rv0678f115-1 (F) | **ACCCAACTGAATGGAGC**TGTTGGGCTGGCTGCTGGTGTGTGAT |
| Rv0678r336-1 (R) | **ACGCACTTGACTTGTCTTC**GGCCATTGCCCGGATGCGTTCA |
| Rv0678f115-2 (F) | **ACCCAACTGAATGGAGC**TATTGGGCTGGtTGCTGGTGTGTGAT |
| Rv0678r336-2 (R) | **ACGCACTTGACTTGTCTTC**GGCCATCGCCCGGATGCGCTCA |
| Rv0678f291-1 (F) | **ACCCAACTGAATGGAGC**AACGCTTTCGCGGCTGGCGAG |
| Rv0678f291-2 (F) | **ACCCAACTGAATGGAGC**AATGCTTTCGCGGCCGGCGAG |
| Rv0678r+24 (R) | **ACGCACTTGACTTGTCTTC**TCGGTCAGATTGCGAGGTTGCTCATCA |
| ***pepQ* (1119 bp)** -33 bp upstream of gene to 1112 bp in gene** | |
| pepQf-60 (F) | **ACCCAACTGAATGGAGC**CAACCCGCGCAGCATCCAGTTAGTCAT |
| pepQr152 (R) | **ACGCACTTGACTTGTCTTC**CGCGCTCATCGGCGAACACCA |
| pepQf199 (F) | **ACCCAACTGAATGGAGC**AAGCGCCCGACCTCGAAGTGGC |
| pepQr421 (R) | **ACGCACTTGACTTGTCTTC**CCAGCTCGCCGGCGTCTTTAACCT |
| pepQf496 (F) | **ACCCAACTGAATGGAGC**GCCGAACCGAACGGCAGGTGAGC |
| pepQr713-1 (R) | **ACGCACTTGACTTGTCTTC**ACGAAGGTGCGGGTCATATCGGAGTGGTA |
| pepQr713-2 (R) | **ACGCACTTGACTTGTCTTC**ACGAAGGTGTGGGTCATATCGCAGTGGTA |
| pepQf899 (F) | **ACCCAACTGAATGGAGC**GCAGATACATGAAGCGCCGGGCATC |
| pepQr+19 (R) | **ACGCACTTGACTTGTCTTC**TGGTCGCCACGTGGGTCTCCTACAGA |
| pepQf74 (F) | **ACCCAACTGAATGGAGC**CGACCTGATAAACGTGCGATATCTATCAGGCTTC |
| pepQr324 (R) | **ACGCACTTGACTTGTCTTC**GTCCAGGCCGTCCACCGTGACCA |
| pepQf351 (F) | **ACCCAACTGAATGGAGC**AACACCGAGTTGGTGCGGGCATCC |
| pepQr535 (R) | **ACGCACTTGACTTGTCTTC**GGGCCTCCAGCTCGCGGCTCAC |
| pepQf640 (F) | **ACCCAACTGAATGGAGC**CGGCGATTTCGTGAAGATCGACTTCGG |
| pepQr968 (R) | **ACGCACTTGACTTGTCTTC**GTCACCACGGAGCCCGCCAGTAGTGTA |

**Supplementary Table 2.**

**Proportion of patients on each TB drug (other than BDQ) and the drugs' likely effectiveness based on WGS, stratified by follow-up phenotypic BDQ result.** No single drug was more likely to be used in BDQ-susceptible or -resistant patients, however, in patients receiving LFX, LFX was more likely to be ineffectively used in BDQ-resistant than -susceptible patients. The same applied to PZA and CFZ. This highlights the need for DST to prevent likely ineffective treatment and alternative drugs. Data are n/N (%). Abbreviations: BDQ – bedaquiline, CFZ – clofazimine, DST – drug susceptibility testing, LFX – levofloxacin, PZA – pDST – phenotypic drug susceptibility testing, pyrazinamide, TB – tuberculosis, WGS – whole genome sequencing.

| | **Overall (n=38)** | **Follow-up BDQ pDST status** | |
|---|---|---|---|
| | | **Susceptible (n=16)** | **Resistant (n=22)** |
| **High dose isoniazid**[†] | 21/38 (55) | 9/16 (56) | 12/22 (55) p=0.917 |
| **Likely effective** | 20/20 (100) | 8/8 (100) | 12/12 (100) p>0.999 |
| **Pyrazinamide** | 35/38 (92) | 15/16 (94) | 20/22 (91) p=0.749 |
| **Likely effective** | 11/3 (33) | 8/14 (57) | 3/19 (16) **p=0.013** |
| **Ethambutol** | 25/38 (66) | 13/16 (81) | 12/22 (55) p=0.087 |
| **Likely effective** | 6/37 (16) | 5/13 (38) | 1/11 (9) p=0.098 |
| **Levofloxacin** | 25/38 (66) | 11/16 (69) | 14/22 (64) p=0.743 |
| **Likely effective** | 4/24 (17) | 4/10[*] (40) | 0/14 (0) **p=0.010** |
| **Moxifloxacin** | 24/38 (63) | 11/16 (69) | 13/22 (59) p=0.542 |
| **Likely effective** | 5/23 (22) | 3/11 (27) | 2/12 (17) p=0.538 |
| **Capreomycin** | 1/38 (3) | 1/16 (6) | 0/22 (0) p=0.235 |
| **Likely effective** | 1/1 (100) | 1/1 (100) | N/A |
| **Amikacin** | 3/38 (8) | 1/16 (6) | 2/22 (9) p=0.749 |
| **Likely effective** | 2/3 (67) | 1/1 (100) | 1/2 (50) p=0.387 |
| **Kanamycin** | 20/38 (53) | 8/16 (50) | 12/22 (55) p=0.782 |
| **Likely effective** | 14/19 (74) | 7/8 (88) | 7/11 (64) p=0.244 |

| | | | |
|---|---|---|---|
| **Linezolid** | 25/38<br>(66) | 10/16<br>(63) | 15/22<br>(68)<br>p=0.716 |
| **Likely effective** | 23/24<br>(96) | 9/9<br>(100) | 14/15<br>(93)<br>p=0.429 |
| **Clofazimine** | 25/38<br>(66) | 8/16<br>(50) | 17/22<br>(77)<br>p=0.080 |
| **Likely effective** | 8/36<br>(22) | 6/15<br>(40) | 2/21<br>(10)<br>**p=0.030** |
| **Terizidone** | 33/38<br>(87) | 14/16<br>(88) | 19/22<br>(86)<br>p=0.919 |
| **Likely effective** | 28/31<br>(90) | 13/13<br>(100) | 15/18<br>(83)<br>p=0.121 |
| **Delamanid** | 9/38<br>(34) | 3/16<br>(19) | 6/22<br>(27)<br>p=0.542 |
| **Likely effective** | 9/9<br>(100) | 3/3<br>(100) | 6/6<br>(100)<br>p>0.999 |
| **Ethionamide** | 26/38<br>(68) | 11/16<br>(69) | 15/22<br>(68)<br>p=0.970 |
| **Likely effective** | 6/25<br>(24) | 4/11<br>(36) | 2/13<br>(15)<br>p=0.237 |
| **4-aminosalicylic acid** | 21/38<br>(55) | 7/16<br>(44) | 14/22<br>(64)<br>p=0.224 |
| **Likely effective** | 18/19<br>(95) | 5/6<br>(83) | 13/13<br>(100)<br>p=0.131 |

Two patients excluded due to unknown treatment regimens
Four WGS results unavailable
*10 mg/kg per South African treatment guideline (12)

**Supplementary Table 3.**

**Select characteristics of patients based on follow-up BDQ resistance status after baseline BDQ resistant strains and, separately, also reinfections were excluded.** Compared to comparisons involving all patients (**Main text, Results**), done without exclusions based on baseline BDQ phenotype and reinfection, fewer variables were significantly associated with resistance at follow-up when both these exclusions were made (only ≤4 likely effective drugs). Data are n/N or median (IQR) unless otherwise stated. Abbreviations: BDQ – bedaquiline, CFZ – clofazimine, FQ – fluoroquinolone, IQR – interquartile range, OR – odds ratio, R – resistance, TB – tuberculosis, WGS – whole genome sequencing

| | Baseline BDQ R omitted | | | Baseline BDQ R and reinfections omitted | | |
|---|---|---|---|---|---|---|
| | BDQ phenotype at follow-up | | | BDQ phenotype at follow-up | | |
| | Susceptible (n=18) | Resistant (n=19) | OR (95% CI) | Susceptible (n=13) | Resistant (n=18) | OR (95% CI) |
| **Baseline FQ resistance*** | 4/18 (22) | 12/18 (67) **p=0.01** | 7 (2-31) **p=0.01** | 4/13 (31) | 11/17 (65) p=0.07 | 4 (1-19) p=0.06 |
| **Any CFZ exposure (prior or concurrent)** | 10/18 (56) | 16/19 (84) p=0.06 | 4 (1-20) p=0.05 | 8/13 (62) | 15/18 (83) p=0.17 | 3 (0.6-17) p=0.17 |
| **≤4 likely effective drugs (excluding BDQ)†** | 5/16 (31) | 16/18 (89) **p<0.01** | 16 (3-99) **p<0.01** | 3/11 (27) | 15/17 (88) **p<0.01** | 20 (2-145) **p<0.01** |
| **Unfavourable outcome** | 6/18 (33) | 17/19 (89) **p<0.01** | Not calculable (few resistance cases with favourable outcomes) | 5/13 (38) | 16/18 (89) **p<0.01** | Not calculable (few resistance cases with favourable outcomes) |

*Detected by WGS or programmatic line probe assay. One result unavailable

†Two patients were excluded due to unknown background TB drug regimens and two WGS sequencing results unavailable

**Supplementary Table 4.**

**Phenotypic and genotypic DST result for three patients with phenotypic BDQ resistance at baseline (all also resistant at follow-up).** TDS frequently detected variants that WGS did not, however, one patient at follow-up (37-B-08) had a variant (*pepQ* 693 A ins) exclusively detected by WGS. The last row shows summary data. As detailed in **Methods**, TDS was done on *Rv0678*, *atpE*, and *pepQ*. WGS was also done on at *Rv0676c*, *Rv0677c* and *Rv1979c*. No variants were detected in *atpE*, *Rv0677c* and *Rv1979c* and these columns are omitted. There was no evidence of reinfection. Prior CFZ exposure, days on BDQ (with programmatic treatment outcome), SNP distances, and the specific variant (proportion of reads indicated) are shown (blue indicates variant loss, red indicates gain). Variants previously described (13) are bolded. The only variants the baseline BDQ-resistant patient 26-A26 had were in *Rv0676c*, however, these also appeared in BDQ-susceptible isolates **(Supplementary Table 5)**, suggesting these are lineage markers and this isolate has an unknown resistance mechanism that, at follow-up, led to the emergence of previously described *Rv0678* variants. Abbreviations: BDQ – bedaquiline, CFZ – clofazimine, indels – insertions and deletions, R – resistant, S – susceptible, SNPs – single nucleotide polymorphisms, TDS – targeted deep sequencing, WGS – whole genome sequencing, WT – wildtype. Data are % unless otherwise stated.

| **Patient identifier (CFZ exposure)** | **Days BDQ treatment duration (treatment outcome)** | **Timepoint** | **Phenotypic DST** | **SNP distance (interpretation)** | **TDS** | | **WGS** | | |
|---|---|---|---|---|---|---|---|---|---|
| | | | | | ***Rv0678*** | ***pepQ*** | ***Rv0678*** | ***pepQ*** | ***Rv0676c*** |
| 37-B08<br>(Yes) | 208<br>(Cured) | Baseline | R | 7<br>(likely intrapatient evolution) | *SNPs*<br>73 G/A (2)<br>119 T/C (9)<br>122 G/T (2)<br>194 G/T (1)<br>226 C/T (19)<br>391 C/T (2)<br>417 G/T (2)<br>*Indels*<br>**192 G ins (1)**<br>141 T del (2)<br>482 C del (10) | WT (100) | *SNPs*<br>226 C/T (26) | WT (100) | *SNPs*<br>1266 G/C (22)<br>2842 T/C (100) |
| | | Follow-up | R | | WT (100) | WT (100) | WT (100) | *Indels*<br>693 A ins (97) | *SNPs*<br>2842 T/C (100) |
| 26-A26*<br>(Yes) | 201<br>(Treatment failed) | Baseline | R | 0<br>(no change) | *SNPs*<br>-11 C/A (100) | WT (100) | *SNPs*<br>-11 C/A (100) | WT (100) | *SNPs*<br>**2299 C/T (100)**<br>**2381 G/A (100)**<br>**2842 T/C (100)** |
| | | Follow-up | R | | *SNPs*<br>-11 C/A (100)<br>394 C/T (3)<br>461 T/C (17)<br>*Indels*<br>132 GT ins (10)<br>137 TGA ins (3) | WT (100) | *SNPs*<br>-11 C/A (98)<br>461 T/C (21)<br>*Indels*<br>132 GT ins (13)<br>**192 G ins (50)** | WT (100) | *SNPs*<br>**2299 C/T (100)**<br>**2381 G/A (100)**<br>**2842 T/C (100)** |

| Patient identifier (CFZ exposure) | Days BDQ treatment duration (treatment outcome) | Timepoint | Phenotypic DST | SNP distance (interpretation) | TDS | | | | WGS | | | | | | | | |
|---|---|---|---|---|---|---|---|---|---|---|---|---|---|---|---|---|---|
| | | | | | *Rv0678* | | | *pepQ* | *Rv0678* | | | *pepQ* | | | *Rv0676c* | | |
| | | | | | **192 G ins (57)**<br>217 A del (2)<br>218 T del (2) | | | | | | | | | | | | |
| 29-A29<br>(Yes) | 91<br>(Died) | Baseline | R | 0<br>(no change) | *SNPs*<br>**136 T/C (100)** | | | WT<br>(100) | *SNPs*<br>**136 T/C (100)** | | | WT (100) | | | *SNPs*<br>**2381 G/A (100)**<br>**2842 T/C (100)** | | |
| | | Follow-up | R | | *SNPs*<br>**136 T/C (100)** | | | WT<br>(100) | *SNPs*<br>**136 T/C (100)** | | | WT (100) | | | *SNPs*<br>**2381 G/A (100)**<br>**2842 T/C (100)** | | |
| **Sub-total<br>(Yes=3)** | **Median (IQR) days of BDQ treatment**<br><br>201 (91-208)<br><br>Cured (n=1)<br><br>Treatment failed (n=1)<br><br>Died (n=1) | Baseline | R<br>n=3 | **Median (IQR) variant distance**<br><br>0 (0-7)<br><br>Likely intrapatient evolution (n=1)<br><br>No change (n=2) | *SNPs*<br>-11 C/A<br>73 G/A<br>119 T/C<br>122 G/T<br>**136 T/C**<br>194 G/T<br>226 C/T<br>391 C/T<br>417 G/T<br>*Indels*<br>**192 G ins**<br>141 T del<br>482 C del | *n*<br>1<br>1<br>1<br>1<br>**1**<br>1<br>1<br>1<br>1<br>*n*<br>**1**<br>1<br>1 | *Median (IQR)*<br>N/A<br>N/A<br>N/A<br>N/A<br>**N/A**<br>N/A<br>N/A<br>N/A<br>N/A<br><br>**N/A**<br>N/A<br>N/A | - | *SNPs*<br>-11 C/A<br>**136 T/C**<br>226 C/T | *n*<br>1<br>**1**<br>1 | *Median (IQR)*<br>N/A<br>N/A<br>N/A | - | | | *SNPs*<br>1266 G/C<br>**2299 C/T**<br>**2381 G/A**<br>**2842 T/C** | *n*<br>1<br>**1**<br>**2**<br>**3** | *Median (IQR)*<br>N/A<br>**N/A**<br>**100 (100-100)**<br>**100 (100-100)** |
| | | Follow-up | R<br>n=3 | | *SNPs*<br>-11 C/A<br>**136 T/C**<br>394 C/T<br>461 T/C<br>*Indels*<br>132 GT ins<br>137 TGA ins<br>**192 G ins**<br>217 A del<br>218 T del | *n*<br>1<br>**1**<br>1<br>1<br>*n*<br>1<br>1<br>**1**<br>1<br>1 | *Median (IQR)*<br>N/A<br>**N/A**<br>N/A<br>N/A<br><br>N/A<br>N/A<br>**N/A**<br>N/A<br>N/A | - | *SNPs*<br>-11 C/A<br>**136 T/C**<br>461 T/C<br>*Indels*<br>132 GT ins<br>**192 G ins** | *n*<br>1<br>**1**<br>1<br>*n*<br>1<br>**1** | *Median (IQR)*<br>N/A<br>**N/A**<br>N/A<br><br>N/A<br>**N/A** | *Indels*<br>693 A ins | *n*<br>1 | *Median (IQR)*<br>N/A | *SNPs*<br>**2299 C/T**<br>**2381 G/A**<br>**2842 T/C** | *n*<br>**1**<br>**2**<br>**3** | *Median (IQR)*<br>**N/A**<br>**100 (100-100)**<br>**100 (100-100)** |

75

**Supplementary Table 5.**

**Phenotypic and genotypic DST result for patients with no phenotypic BDQ resistance and at baseline and follow-up.** The last row shows summary data. As detailed in **Methods**, TDS was done on *Rv0678*, *atpE*, and *pepQ*. WGS was also done on at *Rv0676c*, *Rv0677c* and *Rv1979c*. No variants were detected in *atpE*, and *Rv0677c* and these columns are omitted. When comparing baseline and follow-up isolates, 63% (10/16) with newly gained resistance appeared to be due to intrapatient evolution, however, 31% (5/16) patients had evidence of reinfection. Prior CFZ exposure, days on BDQ (with programmatic treatment outcome), SNP distances, and the specific variant (proportion of reads indicated) are shown (blue indicates variant loss, red indicates gain). Variants previously described (13) are bolded. In baseline isolates, 35% (6/17) compared to 20% (3/15) follow up isolates had *Rv0678* -11 C/A variants detected. Two phenotypically susceptible isolates have *pepQ* variants detected (32-B03, 13-A13). Abbreviations: BDQ – bedaquiline, CFZ – clofazimine, indels – insertions and deletions, R – resistant, S – susceptible, SNPs – single nucleotide polymorphisms, TDS – targeted deep sequencing, WGS – whole genome sequencing, WT – wildtype. Data are % unless otherwise stated.

| **Patient identifier (CFZ exposure)** | **Days of BDQ treatment duration (treatment outcome)** | **Timepoint** | **Phenotypic DST** | **SNP distance (interpretation)** | **TDS** | | **WGS** | | | |
|---|---|---|---|---|---|---|---|---|---|---|
| | | | | | ***Rv0678*** | ***pepQ*** | ***Rv0678*** | ***pepQ*** | ***Rv0676c*** | ***Rv1979c*** |
| 31-B02<br>(Yes) | 862<br>(Cured) | Baseline | S | 3<br>(likely intrapatient evolution) | WT (100) | WT (100) | WT (100) | WT (100) | *SNPs*<br>**2842 T/C (100)** | WT (100) |
| | | Follow-up | S | | WT (100) | WT (100) | WT (100) | WT (100) | *SNPs*<br>**2842 T/C (100)** | WT (100) |
| 06-A06<br>(Yes) | 178<br>(Treatment completed) | Baseline | S | 2<br>(likely intrapatient evolution) | WT (100) | WT (100) | WT (100) | WT (100) | *SNPs*<br>**2299 C/T (100)**<br>**2381 G/A (100)**<br>**2842 T/C (100)**<br>2889 C/T (100) | WT (100) |
| | | Follow-up | S | | WT (100) | WT (100) | WT (100) | WT (100) | *SNPs*<br>**2299 C/T (100)**<br>**2381 G/A (100)**<br>**2842 T/C (100)**<br>2889 C/T (100) | WT (100) |
| 15-A15<br>(Yes) | 168<br>(Treatment completed) | Baseline | S | 838<br>(likely reinfection) | WT (100) | WT (100) | WT (100) | WT (100) | *SNPs*<br>1792 A/T (100)<br>**2842 T/C (100)** | WT (100) |
| | | Follow-up | S | | WT (100) | WT (100) | WT (100) | WT (100) | *SNPs*<br>**2842 T/C (100)** | WT (100) |
| 02-A02<br>(Yes) | 186<br>(Died) | Baseline | S | 4<br>(likely intrapatient evolution) | *SNPs*<br>-11 C/A (100) | WT (100) | *SNPs*<br>-11 C/A (100) | WT (100) | *SNPs*<br>**2381 G/A (100)**<br>**2842 T/C (100)**<br>**2889 C/T (100)** | WT (100) |
| | | Follow-up | S | | *SNPs*<br>-11 C/A (100) | WT (100) | *SNPs*<br>-11 C/A (100) | WT (100) | *SNPs*<br>**2381 G/A (100)**<br>**2842 T/C (100)**<br>**2889 C/T (100)** | WT (100) |
| 05-A05<br>(Yes) | 343<br>(Died) | Baseline | S | 2<br>(likely intrapatient evolution) | **SNPs**<br>-11 C/A (100) | WT (100) | *SNPs*<br>-11 C/A (99) | WT (100) | *SNPs*<br>**2299 C/T (100)**<br>**2381 G/A (100)** | WT (100) |

| Patient identifier (CFZ exposure) | Days of BDQ treatment duration (treatment outcome) | Timepoint | Phenotypic DST | SNP distance (interpretation) | TDS | | WGS | | | |
|---|---|---|---|---|---|---|---|---|---|---|
| | | | | | *Rv0678* | *pepQ* | *Rv0678* | *pepQ* | *Rv0676c* | *Rv1979c* |
| | | | | | | | | | **2842 T/C (100)** | |
| | | Follow-up | S | | **SNPs**<br>-11 C/A (100) | WT (100) | *SNPs*<br>-11 C/A (100) | WT (100) | *SNPs*<br>**2299 C/T (100)**<br>**2381 G/A (100)**<br>**2842 T/C (100)** | WT (100) |
| 32-B03<br>(Yes) | 126<br>(Died) | Baseline | S | 5<br>(likely intrapatient evolution) | WT (100) | WT (100) | WT (100) | WT (100) | *SNPs*<br>**2381 G/A (100)**<br>**2842 T/C (100)** | *SNPs*<br>**1266 C/T (100)** |
| | | Follow-up | S | | WT (100) | *Indels*<br>241 G ins (100)<br>219 G del (100) | WT (100) | *Indels*<br>241 G in (98)<br>219 G del (100) | *SNPs*<br>**2381 G/A (100)**<br>**2842 T/C (100)** | *SNPs*<br>**1266 C/T (100)** |
| 11-A11<br>(Yes) | 218<br>(Loss to follow-up) | Baseline | S | 77<br>(likely reinfection) | WT (100) | WT (100) | WT (100) | WT (100) | *SNPs*<br>**2842 T/C (100)** | WT (100) |
| | | Follow-up | S | | WT (100) | WT (100) | WT (100) | WT (100) | *SNPs*<br>**2842 T/C (100)** | WT (100) |
| 24-A24<br>(Yes) | 168<br>(Loss to follow-up) | Baseline | S | Noncalculable | *SNPs*<br>-11 C/A (100) | WT (100) | *SNPs*<br>-11 C/A (99) | WT (100) | *SNPs*<br>**2299 C/T (100)**<br>**2381 G/A (100)**<br>**2842 T/C (100)** | WT (100) |
| | | Follow-up | S | | *SNPs*<br>-11 C/A (100) | WT (100) | No WGS available | | | |
| 17-A17<br>(Yes) | 283<br>(Not evaluated) | Baseline | S | 2<br>(likely intrapatient evolution) | WT (100) | WT (100) | WT (100) | WT (100) | *SNPs*<br>**2299 C/T (100)**<br>**2381 G/A (100)**<br>**2842 T/C (100)** | *SNPs*<br>151 A/T (100) |
| | | Follow-up | S | | WT (100) | WT (100) | WT (100) | WT (100) | *SNPs*<br>**2299 C/T (100)**<br>**2381 G/A (100)**<br>**2842 T/C (100)** | *SNPs*<br>151 A/T (100) |
| 20-A20<br>(No) | 168<br>(Cured) | Baseline | S | 240<br>(likely reinfection) | *SNPs*<br>-11 C/A (100) | WT (100) | *SNPs*<br>- 11 C/A (100) | WT (100) | *SNPs*<br>**2299 C/T (100)**<br>**2381 G/A (100)**<br>**2842 T/C (100)** | WT (100) |
| | | Follow-up | S | | WT (100) | WT (100) | WT (100) | WT (100) | *SNPs*<br>**2299 C/T (100)**<br>**2381 G/A (100)**<br>**2842 T/C (100)** | WT (100) |
| 30-B01<br>(No) | 843<br>(Cured) | Baseline | S | 0<br>(no change) | WT (100) | WT (100) | WT (100) | WT (100) | *SNPs*<br>**2842 T/C (100)** | WT (100) |
| | | Follow-up | S | | WT (100) | WT (100) | WT (100) | WT (100) | *SNPs*<br>**2842 T/C (100)** | WT (100) |
| 08-A08<br>(No) | 168<br>(Treatment completed) | Baseline | S | Noncalculable | *SNPs*<br>-11 C/A (100) | WT (100) | *SNPs*<br>-11 C/A (99) | WT (100) | *SNPs*<br>**2299 C/T (100)**<br>**2381 G/A (100)**<br>**2842 T/C (100)** | WT (100) |
| | | Follow-up | S | | WT (100) | WT (100) | No WGS available | | | |
| 16-A16<br>(No) | 170<br>(Treatment completed) | Baseline | S | 1245<br>(likely reinfection) | WT (100) | WT (100) | WT (100) | WT (100) | *SNPs*<br>**2842 T/C (100)** | WT (100) |
| | | Follow-up | S | | WT (100) | WT (100) | WT (100) | WT (100) | *SNPs*<br>**2299 C/T (100)**<br>**2381 G/A (100)** | *SNPs*<br>151 A/T (100) |

| **Patient identifier (CFZ exposure)** | **Days of BDQ treatment duration (treatment outcome)** | **Timepoint** | **Phenotypic DST** | **SNP distance (interpretation)** | **TDS** | | **WGS** | | | |
|---|---|---|---|---|---|---|---|---|---|---|
| | | | | | ***Rv0678*** | ***pepQ*** | ***Rv0678*** | ***pepQ*** | ***Rv0676c*** | ***Rv1979c*** |
| | | | | | | | | | **2842 T/C (100)** | |
| 01-A01<br>(No) | 234<br>(Died) | Baseline | S | 2<br>(likely intrapatient evolution) | *SNPs*<br>-11 C/A (100) | WT (100) | *SNPs*<br>-11 C/A (100) | WT (100) | *SNPs*<br>**2299 C/T (100)**<br>**2381 G/A (100)**<br>**2842 T/C (100)** | WT (100) |
| | | Follow-up | S | | *SNPs*<br>-11 C/A (100) | WT (100) | *SNPs*<br>-11 C/A (100) | WT (100) | *SNPs*<br>**2299 C/T (100)**<br>**2381 G/A (100)**<br>**2842 T/C (100)** | WT (100) |
| 07-A07<br>(No) | 168<br>(Died) | Baseline | S | 1271<br>(likely reinfection) | WT (100) | WT (100) | WT (100) | WT (100) | *SNPs*<br>**2299 C/T (100)**<br>**2381 G/A (100)**<br>**2842 T/C (100)**<br>2889 C/T (100) | WT (100) |
| | | Follow-up | S | | WT (100) | WT (100) | WT (100) | WT (100) | *SNPs*<br>**2842 T/C (100)** | WT (100) |
| 23-A23<br>(No) | 168<br>(Died) | Baseline | S | 4<br>(likely intrapatient evolution) | WT (100) | WT (100) | WT (100) | WT (100) | *SNPs*<br>**2381 G/A (100)**<br>**2842 T/C (100)** | *SNPs*<br>**1266 C/T (100)** |
| | | Follow-up | S | | WT (100) | WT (100) | WT (100) | WT (100) | *SNPs*<br>**2381 G/A (100)**<br>**2842 T/C (100)** | *SNPs*<br>**1266 C/T (100)** |
| 34-B05<br>(No) | 276<br>(Loss to follow-up) | Baseline | S | 2<br>(likely intrapatient evolution) | WT (100) | WT (100) | WT (100) | WT (100) | *SNPs*<br>**2299 C/T (100)**<br>**2381 G/A (100)**<br>**2842 T/C (100)** | *SNPs*<br>151 A/T (100) |
| | | Follow-up | S | | WT (100) | WT (100) | WT (100) | WT (100) | *SNPs*<br>**2299 C/T (100)**<br>**2381 G/A (100)**<br>**2842 T/C (100)** | *SNPs*<br>151 A/T (100) |
| 13-A13<br>(No) | 675<br>(Not evaluated) | Baseline | No Result | 5<br>(likely intrapatient evolution) | WT (100) | WT (100) | WT (100) | WT (100) | *SNPs*<br>**2842 T/C (100)** | WT (100) |
| | | Follow-up | S | | WT (100) | WT (100) | WT (100) | *SNPs*<br>260 C/G | *SNPs*<br>**2842 T/C (100)** | WT (100) |
| **(Sub-total (Yes=9 No=9)** | **Median (IQR) days of BDQ treatment**<br>181 (168-267)<br>Cured (n=3)<br>Treatment completed (n=4)<br>Died (n=6)<br>Loss to follow-up (n=3)<br>Not evaluated (n=2) | Baseline | S<br>n=17 | **Median (IQR) variant distance**<br>4 (2-148)<br>No change (n=1)<br>Likely intrapatient evolution (n=10)<br>Likely reinfection (n=5)<br>Noncalculable (n=2) | *SNPs* / *n* / *Median (IQR)*<br>-11 C/A / 6 / 100 (100-100) | - | *SNPs* / *n* / *Median (IQR)*<br>-11 C/A / 6 / 100 (99-100) | - | *SNPs* / *n* / *Median (IQR)*<br>1792 A/T / 1 / N/A<br>**2299 C/T / 8 / 100 (100-100)**<br>**2381 G/A / 12 / 100 (100-100)**<br>**2842 T/C / 18 / 100 (100-100)**<br>2889 C/T / 3 / 100 (100-100) | *SNPs* / *n* / *Median (IQR)*<br>151 A/T / 2 / 100 (100-100)<br>**1266 C/T / 2 / 100 (100-100)** |
| | | Follow-up | S<br>n=18 | | *SNPs* / *n* / *Median (IQR)* | *Indels* / *n* / *Median (IQR)* | *SNPs* / *n* / *Median (IQR)* | *SNPs* / *n* / *Median (IQR)* | *SNPs* / *n* / *Median (IQR)* | *SNPs* / *n* / *Median (IQR)* |

| **Patient identifier (CFZ exposure)** | **Days of BDQ treatment duration (treatment outcome)** | **Timepoint** | **Phenotypic DST** | **SNP distance (interpretation)** | **TDS** | | | | | | **WGS** | | | | | | | | | | | |
|---|---|---|---|---|---|---|---|---|---|---|---|---|---|---|---|---|---|---|---|---|---|---|
| | | | | | ***Rv0678*** | | | ***pepQ*** | | | ***Rv0678*** | | | ***pepQ*** | | | ***Rv0676c*** | | | ***Rv1979c*** | | |
| | | | | | -11 C/A | 4 | 100 (100-100) | 241 G ins | 1 | N/A | -11 C/A | 3 | 100 (100-100) | 260 C/G | 1 | N/A | **2299 C/T** | **6** | **100 (100-100)** | 151 A/T | 3 | 100 (100-100) |
| | | | | | | | | 219 G del | 1 | N/A | | | | *Indels* | *n* | *Median (IQR)* | **2381 G/A** | **10** | **100 (100-100)** | **1266 C/T** | **2** | **100 (100-100)** |
| | | | | | | | | | | | | | | 241 G ins | 1 | N/A | **2842 T/C** | **16** | **100 (100-100)** | | | |
| | | | | | | | | | | | | | | 219 G del | 1 | N/A | | | | | | |
| | | | | | | | | | | | | | | | | | 2889 C/T | 2 | 100 (100-100) | | | |
